# Discriminatory experiences and depression among Chinese lesbian, gay, and bisexual individuals in the United States: A moderated mediation modeling analysis

**DOI:** 10.1101/2024.08.15.24312038

**Authors:** Hui Xie, Binx Yezhe Lin, Xinyi Jiang, Wisteria Deng

## Abstract

Chinese individuals in the U.S. have suffered substantial discrimination during the COVID19 pandemic, a stressor exacerbated by multiple minority identities (e.g., sexual and gender minority). This study aimed to explore the mechanisms of discriminatory experiences, specifically how it interacted with stressors like internalized homophobia and protective factors such as resilience, to affect depressive symptoms among Chinese individuals with multiple minority identities. Between 2022-2023, 272 Chinese LGB individuals residing in the U.S. (Mean_[age]_=28.36; SD=5.01) was surveyed anonymously using the Everyday discrimination Scale, the Internalized Homophobia Scale, the Conner-Davidson Resilience scale, and the Center for Epidemiologic Studies Depression Scale. Moderation and mediation analyses were conducted using STATA 17 and JASP 0.18.3. Replicating the positive association between discriminatory experience and depressive symptoms, we found that discrimination explained depressive symptoms directly and through increasing internalized homophobia. As a protective factor, resilience moderated the internalized homophobia-depressive symptoms link, but not the discrimination-depressive symptoms link. For Chinese LGB individuals with higher levels of resilience, the positive impact of internalized homophobia on depressive symptoms was weaker compared to those with lower resilience. Our findings further the understanding of the mediating and moderating mechanisms between discriminatory experiences and depressive symptoms among individuals with multiple minority identities. Continued research and intervention development on promoting resilience and other protective factors tailored to Chinese LGB individuals in the U.S. are crucial for improving mental health.

## Introduction

The increased discrimination endured by Chinese individuals living in the U.S. since the onset of the COVID-19 pandemic has been well established [1]. Perceived discrimination has been linked with adverse mental and physical health outcomes [2], posing threats to public health and societal cohesion [3]. The Minority Stress Model, a well-established theory that associates stressful minority experiences (e.g., discrimination, internalized stigma and hypervigilance) with adverse health outcomes [4], has helped explain the plight endured by Chinese individuals in the U.S. during this crisis [5]. However, little attention has been directed toward individuals who navigate intersecting minority identities, such as Chinese individuals who also identify as lesbian, gay, bisexual (LGB), or others (henceforth referred to as sexual minorities). This oversight represents a critical gap in the health literature that warrants further exploration.

Unlike the widely accepted Minority Stress Model, theories linking intersecting minority identities and mental health outcomes have been complicated and pointing in diverging directions [6, 7], Some studies have proposed that multiple minority identities (e.g., LGB individuals of color) result in being “multiply marginalized [8, 9],” with each minority identity contributing an additional layer of minority stress. Furthermore, multiple minorities may experience cumulative discrimination and social exclusion, such as racism within the sexual minority community and heterosexism within their racial/ethnic community [10, 11]. These multiple sources of minority stress may decrease the perceived level of social inclusion and safety, leading to chronic vigilance [12] and emotion dysregulation [13].

Conversely, other studies have suggested that multiple minority identities can be a source of resilience [14–16]. Resilience encapsulates the ability to adapt to situational demands and cope with stressors [16]. Minority individuals may benefit from the ability to switch between their multiple identities, which can reduce the essentialization of any one identity group [6]. In addition, the process of developing one’s unique individual identity amidst the flux of multiple cultural backgrounds may promote personal growth and increase self-efficacy in coping with societal stressors [16].

Given the complicated psychological processes underlying multiple minority identities, it is essential to further explore the mediators and moderators along the pathway between minority stressors and adverse mental health outcomes. One such pathway is the association between discrimination and depression [17], which has been particularly highlighted since the COVID-19 pandemic. In the U.S., there has been a rise in discrimination, especially against Chinese and Chinese American individuals since the pandemic [18]. Similarly, more severe depressive symptoms are reported across the population [19], further exacerbated by minority status since the pandemic [20]. To better understand the connection between perceived discrimination and depression among Chinese sexual minority individuals living in the U.S., several mediator and moderator need to be considered.

Among various factors, we hypothesized that internalized homophobia and resilience play crucial roles in linking discrimination and depressive symptoms within this multiple minority group. Based on the psychological mediation framework [13], sexual minority individuals experience increased minority stress, which heightens the risk for negative cognitive processes (e.g., negative thoughts about the self) and subsequent psychopathology (e.g., depression). Such cognitive processes in turn mediate the pathway between minority stressors and psychopathology. In particular, internalized homophobia may reduce individuals’ self-esteem, posing a threat to their view of themselves and the future, thereby increasing their risk for internalizing symptoms such as depression [21]. On the flip side, resilient personality traits may help moderate the link between discrimination and depression [22], especially in multiple minority individuals.

### The present study

Therefore, our work aimed to 1) extend previous studies on examining internalized homophobia as a potential mediator explaining the pathway between discrimination and depression among Chinese sexual minority individuals residing in the U.S.; and 2) explore the moderating effect of resilience on depression. In particular, we investigated the underlying mechanisms linking discrimination and depressive symptoms using a moderated mediation model. This model examined the mediating role of internalized homophobia and the moderating role of resilience on this pathway (Fig 1). We proposed the following hypotheses:

**Figure 1.**
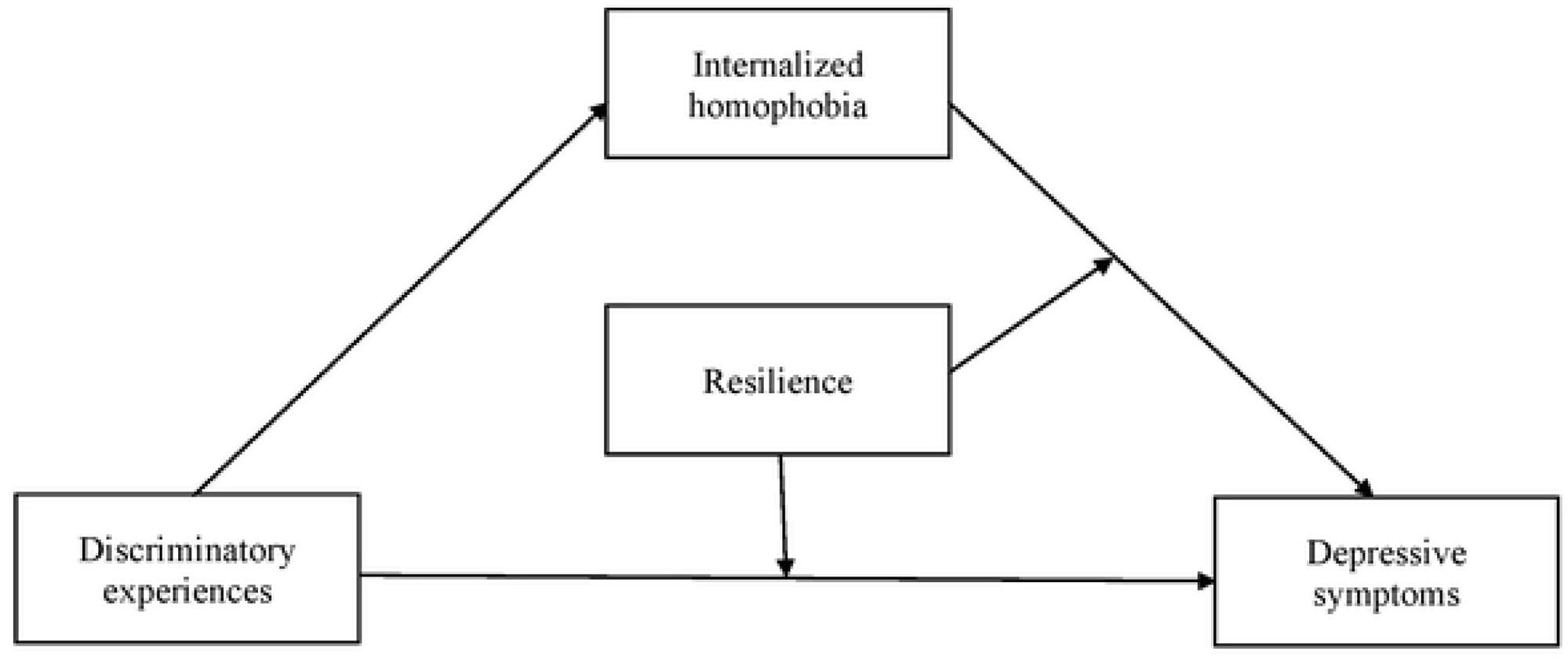
Proposed research model.

Hypothesis 1 (H1): Discriminatory experiences are positively associated with depressive symptoms among Chinese sexual minority individuals living in the U.S.

Hypothesis 2 (H2): Internalized homophobia mediates the relation between discriminatory experiences and depressive symptoms.

Hypothesis 3 (H3): Resilience moderates the association between discriminatory experiences and depressive symptoms, as well as the relationship between internalized homophobia and depressive symptoms.

## Methods

### Participants and recruitment

Between September 2022 and January 2023, a cross-sectional survey data was conducted to assess mental health and overall quality of life in Chinese sexual and/or gender minority (SGM) individuals residing in the U.S. All participants indicated electronic consents over a secure online platform. The term SGM encompassed individuals self-identifying as lesbian, gay, bisexual, asexual, transgender, intersex, etc. Survey recruitment took place through various private chat groups/social media platforms hosted by the China Rainbow Network (CRN), a non-profit organization dedicated to serving and advocating for Chinese LGBT individuals in North America (https://www.crn.ngo/zh-eng/homepage). Inclusion criteria for the survey participants included: 1) age 18-45 years; 2) born in mainland China, Hongkong, or Macau); 3) current residency in the U.S.; 4) self-identification as a SGM individual; and 5) proficiency in English (both reading and speaking). This study was one of the sub-research studies addressing mental health among cisgender, sexual minority individuals only.

All participants provided informed consent electronically, and participation was voluntary. No identifiable questions were collected. Participants who completed the survey were compensated with a $10 Amazon gift card or the equivalent amount donated to CRN. The research was exempt from IRB review at Carilion Clinic (IRB-22-1558).

### Variable measures

#### Discriminatory experiences

Discriminatory experiences were measured using the 9-item Everyday Discrimination Scale [23], assessing chronic and episodic, yet generally minor, instances of interpersonal discrimination. Participants were prompted to rate their experiences based on statements such as “You are treated with less courtesy than other people are,” with response options ranging from 0 (*Never*) to 5 (*Almost every day*). The sum scores were calculated, where higher scores indicated a heightened frequency of discriminatory experience. EDS has been widely used with high reliability in various populations [24, 25], as well as in Chinese/Chinese Americans [26, 27]. In this study, the internal consistency of the scale was high (Cronbach’s alpha = 0.9305).

#### Internalized homophobia

Internalized homophobia was captured using the 20-item Internalized Homophobia Scale developed by Wagner [28] to assess how much an individual’s self-image and identity have been influenced by the internalization of anti-homosexual attitudes and beliefs (e.g., *I wish I were heterosexual*). Participants indicated how much they agreed with each item by using a 5-point Likert scale from 1 (*Strongly disagree*) to 5 (*Strongly agree*). Higher scores indicated higher levels of internalized homophobia. This scale was proven to be applicable for various LGB communities [29, 30]. The internal consistency of the IH scale was high in this study (Cronbach’s alpha = 0.9041).

#### Resilience

Resilience was assessed using a unidimensional self-reported scale from Conner-Davidson Resilience scale-10 items [31], capturing how well one could bounce back from adversity. A sample item is “I am able to adapt when changes occur” with 5-point Liker scale ranging from 0 (*strongly disagree*) to 4 (*strongly agree*). Sum scores were used with higher scores indicating higher levels of resilience. This scale was proven to be applicable for Chinese population [32, 33]. In our study, the internal consistency of the CD-RISC-10 scale was high (Cronbach’s alpha = 0.9345).

#### Depressive symptoms

Depressive symptoms were measured using the 20-item Center for Epidemiologic Studies Depression Scale [34], which assesses the degree of depression present in an individual. A sample item was that “I was bothered by things that usually don’t bother me” with responses ranging from 0 (*Rarely or none of the time/less than 1 day*) to 4 (*Most or all of the time/5-7 days*). A sum scores of 80 or higher was used to define depression. This scale was proven to be applicable for Chinese populations [35, 36]. The internal consistency of the CES-D scale was high in our study (Cronbach’s alpha = 0.9624).

#### Demographic variables

Demographic variables were included age, sex (assigned male at birth, assigned female at birth, or intersex conditions), sexual orientation (gay/lesbian/homosexual, bi/pan-sexual, or other), household income (<$34,999, $35,000-$49,999; $50,000-$74,999, $75,000-$99,999; $100,000-$149,999, or $150,000 and above), and length of staying in the U.S. (less than a year, 1-3 years, 3-5 years, or 5 years and above).

### Statistical analysis

STATA 18 and JASP 0.18.1 were performed to analyze the data. Descriptive statistics was conducted to present demographic characteristics of the sample. Pearson’s correlation analysis was applied to examine the relationships among discriminatory experiences, internalized homophobia, resilience, and depressive symptoms. Mediation modeling analysis was computed, where discriminatory experiences was a predictor, internalized homophobia was a mediator, and depressive symptoms was the response variable using JASP mediation analysis [37] based on lavaan software [38]. Models were estimated by applying bootstrapping and the confidence intervals were calculated using the bias-corrected percentile method, following Biesanz, Falk & Savalei [39]. Next, moderated mediation modeling analysis was used to test moderated effect of resilience on the proposed mediation model above. In the moderated mediation model, all variables were mean-centered prior to the analysis. Age, sex, household income, and length in the U.S. were controlled in both mediation and moderated mediation analyses. The model fit index included root mean square error of approximation (RMSEA), standardized root mean square residual (SRMR), goodness of fit index (GFI) and comparative fit index (CFI).

## Result

### Common-method variance (CMV) test

An explanatory factor analysis (EFA) including all variables using unrotated principal components factor analysis was performed to statistically verify the presence of CMV. The results revealed that 11 factors had eigenvalues >1, and the general factor accounted for only 18.53% of the total variance, which did not exceed the critical value of 40%. It concluded that CMV was not a concern.

### Demographic characteristics

Table 1 presents the demographic characteristics of a sample of 272 Chinese cisgender sexual minority individuals living in the U.S (Mean [age]=28.36; standardized deviation [SD]=5.01). Approximately 44.44% and 59.56% of the sample were AMAB and AFAB, respectively. Almost 72.06% identified as lesbian/gay/homosexual. In terms of household income, a balanced distribution was observed, with 23.16% reporting incomes less than $34,999, 14.71% reporting $50,000-74,999, and 24.63% reporting $150,000 and above. More than 62.00% reporting having lived in the U.S. at least 5 years.

**Table 1.**
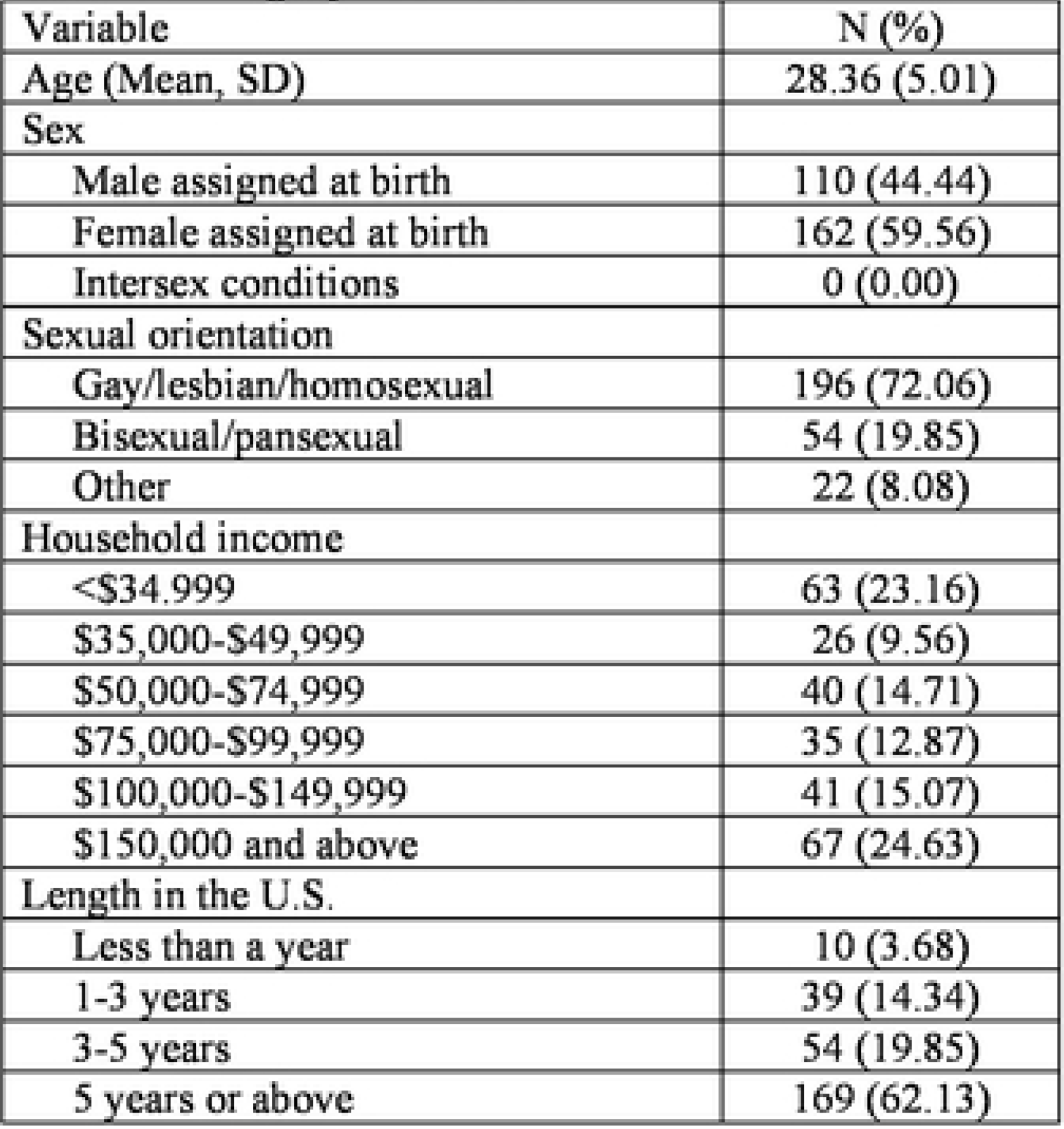
Demoeranhit statistics in Chinese LGB individuals living in the U.S. (N=272)

| Variable | N (%) |
| --- | --- |
| Age (Mean, SD) | 28.36 (5.01) |
| Sex |  |
| Male assigned at birth | 110 (44.44) |
| Female assigned at birth | 162 (59.56) |
| Intersex conditions | 0 (0.00) |
| Sexual orientation |  |
| Gay/lesbian/homosexual | 196 (72.06) |
| Bisexual/pansexual | 54 (19.85) |
| Other | 22 (8.08) |
| Household income |  |
| <\$34,999 | 63 (23.16) |
| \$35,000-\$49,999 | 26 (9.56) |
| \$50,000-\$74,999 | 40 (14.71) |
| \$75,000-\$99,999 | 35 (12.87) |
| \$100,000-\$149,999 | 41 (15.07) |
| \$150,000 and above | 67 (24.63) |
| Length in the U.S. |  |
| Less than a year | 10 (3.68) |
| 1-3 years | 39 (14.34) |
| 3-5 years | 54 (19.85) |
| 5 years or above | 169 (62.13) |

### Bivariate correlations

Table 2 presents the means, SD, and correlations between discriminatory experiences, internalized homophobia, resilience, and depressive symptoms. The results indicated that discriminatory experiences were positively associated with internalized homophobia (r = 0.234; *p* < 0.001) and depressive symptoms (r = 0.398; *p* < 0.001). And internalized homophobia was positively associated with depressive symptoms (r = 0.281; *p* < 0.001). In addition, both discriminatory experiences (r = −0.152; *p* = 0.012), internalized homophobia (r = −0.430; *p* < 0.001), and depressive symptoms (r = −0.316; *p* < 0.001) were negatively associated with resilience.

**Table 2.** Correlations amone variables.

|  | Mean (SD) | 1 | 2 | 3 | 4 | 5 | 6 | 7 | 8 |
| --- | --- | --- | --- | --- | --- | --- | --- | --- | --- |
| 1 Age | 28.36 (5.01) | 1 |  |  |  |  |  |  |  |
| 2 Sex | --- | -0.1338<br>(0.0274) | 1 |  |  |  |  |  |  |
| 3 Household income | --- | 0.3381<br>($<0.001$) | 0.0098<br>(0.8716) | 1 | | | | | |
| 4 Length in the U.S. | --- | 0.3188<br>($<0.001$) | 0.0821<br>(0.1768) | 0.3474<br>($<0.001$) | 1 | | | | |
| 5 Discriminatory experiences | 10.47 (8.48) | -0.0463<br>(0.4469) | 0.0285<br>(0.6401) | -0.0979<br>(0.1071) | -0.0272<br>(0.6551) | 1 |  |  |  |
| 6 Internalized homophobia | 46.33 (13.73) | 0.0278<br>(0.6484) | -0.2203<br>(0.0003) | -0.1441<br>(0.0174) | -0.0759<br>(0.2121) | 0.2338<br>(0.0001) | 1 |  |  |
| 7 Resilience | 24.53 (8.45) | 0.1231<br>(0.0425) | 0.0606<br>(0.3193) | 0.2321<br>(0.0001) | 0.0341<br>(0.5750) | -0.1516<br>(0.0123) | -0.4301<br>($<0.001$) | 1 | |
| 8 Depressive symptoms | 9.49 (5.34) | -0.1537<br>(0.0111) | -0.0537<br>(0.3779) | -0.2628<br>($<0.001$) | -0.2174<br>(0.0003) | 0.3985<br>($<0.001$) | 0.2810<br>($<0.001$) | -0.3164<br>($<0.001$) | 1 |

### Mediation analysis

Table 3 shows the results of mediating effect of internalized homophobia after adjusting for age, sex, household income, length in the U.S. Discriminatory experiences had a significant positive effect on depressive symptoms (β = 0.214; t = 7.285; *p* <0.001), and on internalized homophobia (β = 0.371; t = 4.018; *p* <0.001). When the mediating variable internalized homophobia was added, the direct effect of discriminatory experiences on depressive symptoms was still significant (β = 0.213; t = 6.285; *p* <0.001), indicating partial mediation. Internalized homophobia had a positive effect on depressive symptoms (β = 0.065; t = 3.014; *p* =0.003).

**Table 3.** Testing the mediation effect or discriminatory experienc ondepressive symptoms through internalized homophobia.

|  | Model 1<br>(Depressive symptoms) |  | Model 2 (Internalized<br>homophobia) |  | Model 3 (Depressive<br>symptoms) |  |
| --- | --- | --- | --- | --- | --- | --- |
| | $\beta$ | t | $\beta$ | t | $\beta$ | t |
| Age | -0.021 (0.851) | -0.188 | 0.171 (0.321) | 0.993 | -0.061 (0.322) | -0.991 |
| Sex | 0.467 (0.659) | 0.441 | -6.007 (<0.001) | -3.719*** | -0.242 (0.684) | -0.407 |
| Household income | -0.358 (0.146) | -1.454 | -0.788 (0.036) | -2.092* | -0.331 (0.015) | -2.433* |
| Length in the U.S. | 0.080 (0.902) | 0.123 | -0.415 (0.676) | -0.419 | -0.776 (0.029) | -2.183* |
| Discriminatory experiences | 0.214 (<0.001) | 7.285*** | 0.371 (<0.001) | 4.018*** | 0.213 (<0.001) | 6.285*** |
| Internalized homophobia |  |  |  |  | 0.065 (0.003) | 3.014 |
| R<sup>2</sup> | 0.22 |  | 0.12 |  | 0.27 |  |
| F | 16.12*** |  | 7.51*** |  | 25.31*** |  |
| p | 0.001 |  | 0.001 |  | 0.001 |  |

**Note.** Each column is a regression model that predicts the criterion at the top of the column.
\*\*\* $p < 0.001$ ; \*\* $p < 0.01$ ; \* $p < 0.05$ .

Further bootstrapping results showed internalized homophobia partially mediated the relationship between discriminatory experiences and depressive symptoms, with an indirect effect accounting for 36.7% of the total effect. These findings supported H1 and H2.

### Moderated mediation analysis

The structural equation model regarding the mediating role of internalized homophobia and moderating role of resilience in the relationship between discriminatory experiences and depressive symptom presented a good fit to the data: χ^2^_(13, N = 272)_= 683.473, *p* < 0.001; RSMEA = 0.068; SRMR = 0.052; GFI = 0.970; and CFI = 0.916. In our proposed moderation mediation model, we hypothesized that resilience would moderate direct and indirect effects of internalized homophobia on depressive symptoms in the mediation model. Table 4 shows that results of such moderation mediation analysis using Model 15 on JASP by Hayes. After resilience was put into the mode, the direct effect of discriminatory experiences on depressive symptoms (β = 0.127; t = 1.396; *p* =0.163) and the interaction term between discriminatory experiences and resilience (β = 0.003; t = 0.880; *p* =0.379) were NOT significant (See S1 Table). Meanwhile, the interaction term between internalized homophobia and resilience was significant (β = 0.007; t = 8.928; *p* <0.001). Thus, we adjusted the moderated mediation model using Model 14 on JASP by Hayes. The results show that (Table 4), discriminatory experience had a positive effect on internalized homophobia (β = 0.371; t = 4.018; *p* <0.001), internalized homophobia had negative effect on depressive symptoms (β = −0.164; t = −7.808; *p* <0.001), and discriminatory experience had a positive effect on internalized homophobia (β = 0.202; t = 5.935; *p* <0.001). In addition, the interaction term between internalized homophobia and resilience had a significant impact on depressive symptoms (β = 0.008; t = 9.141; *p* <0.001), indicating that resilience moderated the indirect effect of discriminatory experiences on depressive symptoms via internalized homophobia. This finding partially supported H3.

**Table 4.**
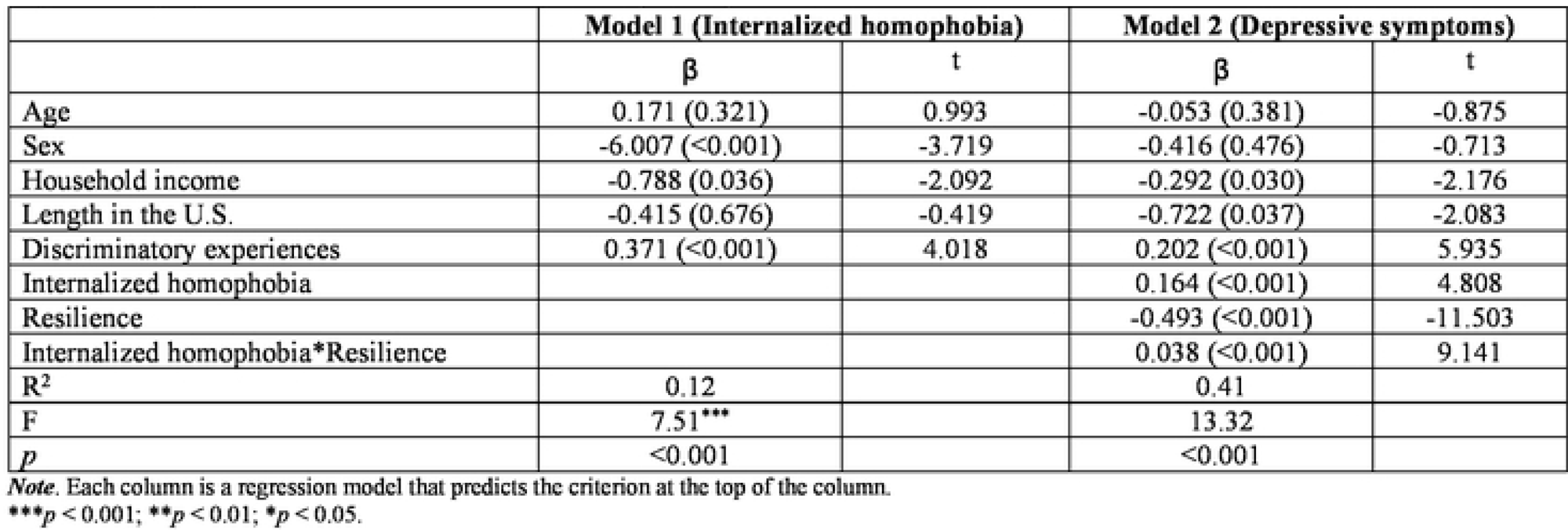
Testing the moderated mediation effect of re.siJience on the relation between discriminatory experiences and deoressive svmntoms *via* internalized homoohobia lN=272)

|  | Model 1 (Internalized homophobia) |  | Model 2 (Depressive symptoms) |  |
| --- | --- | --- | --- | --- |
| | $\beta$ | t | $\beta$ | t |
| Age | 0.171 (0.321) | 0.993 | -0.053 (0.381) | -0.875 |
| Sex | -6.007 (<0.001) | -3.719 | -0.416 (0.476) | -0.713 |
| Household income | -0.788 (0.036) | -2.092 | -0.292 (0.030) | -2.176 |
| Length in the U.S. | -0.415 (0.676) | -0.419 | -0.722 (0.037) | -2.083 |
| Discriminatory experiences | 0.371 (<0.001) | 4.018 | 0.202 (<0.001) | 5.935 |
| Internalized homophobia |  |  | 0.164 (<0.001) | 4.808 |
| Resilience |  |  | -0.493 (<0.001) | -11.503 |
| Internalized homophobia*Resilience |  |  | 0.038 (<0.001) | 9.141 |
| R<sup>2</sup> | 0.12 |  | 0.41 |  |
| F | 7.51*** |  | 13.32 |  |
| p | <0.001 |  | <0.001 |  |

*Note.* Each column is a regression model that predicts the criterion at the top of the column.
\*\*\* $p < 0.001$ ; \*\* $p < 0.01$ ; \* $p < 0.05$ .

A simple slope analysis was conducted to determine the separate relationships between internalized homophobia and depressive symptoms. The results showed that for participants with lower levels of resilience (mean −1SD), internalized homophobia had NOT significant effect on depressive symptoms (slope =0.046, t= 4.558, *p*=0.019). For participants with higher level of resilience (mean +1SD), internalized homophobia had a significant effect on depressive symptoms: simple slope =0.022, t=2.120, *p*=0.034. This indicated that with improvements in resilience among Chinese sexual minority individuals residing in the U.S., the predictive effect of internalized homophobia on depressive symptoms showed a gradual downward trend.

**Table 5.** Conditional indirect effect or internalized homonhobia on denressive svmntoms at snecial level or resilience fN-2721.

| Social support | Mean | Boot indirect effect<br>$\beta$ | Boot SE | p value | Boot 95%CI |
| --- | --- | --- | --- | --- | --- |
| Low<sup>a</sup> | 16.08 | 0.036 | 0.016 | 0.019 | 0.022, 0.051 |
| Moderate<sup>b</sup> | 24.53 | 0.023 | 0.011 | 0.030 | 0.016, 0.044 |
| High<sup>c</sup> | 32.98 | 0.012 | 0.010 | 0.034 | 0.008, 0.029 |

*Note.*
<sup>a</sup>Low was 1 SD below the mean.
<sup>b</sup>Moderate was the mean.
<sup>c</sup>High was 1 SD above the mean.

## Discussion

The significant impact of discriminatory experiences on depressive symptoms has garnered considerable empirical support in previous studies [40, 41]. However, the mechanisms through which these effects occur, particularly the roles of moderating and mediating factors, remain insufficiently understood within this specific population. To address this gap, our study employed a moderated mediation model to examine the interplay between discriminatory experiences, internalized homophobia, and resilience in influencing depressive symptoms. Our findings indicate that internalized homophobia partially mediates the relationship between discriminatory experiences and depressive symptoms. Our moderated mediation model reveals that resilience moderates the relationship between internalized homophobia and depressive symptoms, though it does not moderate the relationship between of discriminatory experiences and internalized homophobia or depressive symptoms. These insights enhance our understanding of the complex pathways linking minority stressors to mental health outcomes and underscore the importance of resilience in mitigating negative effects.

### Discriminatory experiences & depressive symptoms

The present study contributes to existing literature in three significant ways. Firstly, our findings, in accordance with H1, underscore the substantial relationship between discriminatory experiences and depressive symptoms, both directly and indirectly [42, 43]. Despite controlling for socioeconomic factors such as length of time in the U.S. and household income, Chinese sexual minority individuals continue to encounter certain types of discrimination. For instance, Chinese individuals reported experiencing racism/ xenophobia/discrimination/prejudice [44, 45], particularly exacerbated during the pandemic [46, 47]. Also, Chinese MSM individuals report instances of racism in dating (sexual racism) within sexual minority communities [48, 49]. Perceived discrimination across different communities acts as oppressive sources of social discrimination or exclusion in this given population.

In our study, a significant proportion of participants reported experiencing discriminatory incidents at least several times a month. Specially, 22.05% reported “being treated with less courtesy” and 19.40% reported that “people act as if they are better than me”. As a form of microaggression harm [50], whether intentional and unintentional, has a cumulative impact on individuals’ health over time. These subtle forms of discrimination and prejudice carry both informative and meaningful values. Feelings of inferiority, inauthenticity, and social exclusion could have enduring effects on cognitive, emotional, and immunological functioning, even when exposure to minority stress is low [12, 51]. Recipients of microaggressions may feel confused and question these experiences, or they may adapt to them as a way to be “people of color and/or being sexual minorities” in the US. Or some are deprived of the private self presented in everyday life [52]. Eventually, this hinders social connection, inclusion, and protection, which as fundamental constructs of minority stress in relation to adverse mental health.

### Internalized homophobia as a mediator

Consistent with the Psychological Mediation Framework [13] we found supportive evidence for the indirect effect of distal minority stress experiences on depressive symptoms via internalized homophobia (H2). Several studies conducted in Western contexts have examined how internalized homophobia links the experience of distal minority stress and mental health outcomes [53–55]. Another study conducted in Chinese context by Sun et al. [56] better explaining the minority stress processes confronting Chinese men having sex with men. In their framework, stigma internalization was cited as an intermediate mechanism, highlighting the significant impact of norm conformity and family support in this population.

Moving beyond these existing models, our study argues that discriminatory experiences (regardless of sexual orientation, race, nationality, or other minority conditions/identities) act as general oppressive sources that elicit internalized homophobia [57] and emotional dysregulation [13] inwardly. Notably, persistent internalized homophobia may also be correlated to factors such as discrimination salience in family/Chinese culture. Our study shows that only 33.46% reported being out to their parents/family members in our study. Those who have not out to their parents reported significantly higher levels of internalized homophobia than those who came out to their parents (Mean_[IH]_ =48.19 vs. 42.64; *p* =0.0015). The reluctance to come out can be attributed to the negative social climate toward sexual minority individuals arising from cultural norms, conventional marriage ideation, and the one-child policy [56, 58–60]. Many Chinese sexual minority individuals find it necessary to conceal their sexual orientation at home to conform to family expectations and secure financial mobility. Or others find it easier to conceal their sexual orientation than their race/ethnicity when the oppression/discrimination/stigma occurs. Over time, the chronic stress/oppression stemming from those daily life experiences may lead to cognitive dissonance and a denial of sexual identity/internalized racism, as a way to prevent “secondary” harms, resulting in long-term adverse mental health [48, 49, 61].

### Resilience as a moderator

This study makes a unique contribution to the minority stress model by examining the moderation effect of resilience in the pathway between internalized homophobia and depressive symptoms through internalized homophobia (H3). Contrary to our hypothesis, resilience did not moderate the direct effect of discriminatory experiences on depression, aligning with Scandurra et al.’s [62] and contrasting with Li, Yan, Wang et al. [63]. Specially, Chinese sexual minority individuals with higher resilience levels were better able to mitigate the impact of internalized homophobia on depressive symptom compared to those with lower resilience. This supports previous studies highlighting the protective role of resilience in buffering the negative impacts of minority stress on health [53, 64, 65]. Our study also found a positive association between resilience and connectedness to LGB communities, suggesting that resilience can be enhanced through community socialization. Active engagement in the sexual minority community may reduce identity conflicts and thereby alleviating affective symptoms. These findings advocate for strength-based intervention that promote community resilience to address identity conflicts and internalized stigma.

The absence of a significant moderation by resilience on the direct association between discriminatory experiences and depressive symptoms warrants further investigation. One possible explanation is that single discriminatory events can trigger intense and immediate emotion dysregulation, social/interpersonal conflicts, and cognitive disruptions [13], while single kind of resilience or resilience alone may not fully address. For instance, the partial moderating effect might result from a lack of development through family socialization [66]. As many participants’ families might not reside in the U.S., due to their immigrant status, there could be a reluctance to share negative experiences due to emotional restraints and family expectations [52, 66]. Resilience is a multifaceted and context-dependent cognitive development that requires further exploration in populations with multiple minoritized identities.

### Implications

Our findings call to attention the importance of both individual and community resilience [52, 53, 66]. It is also pivotal to promote resilience as related to an individual’s intersecting identities [64]. For instance, integrating resilience with positive coping mechanisms, emotional regulation strategies, and social support for a more comprehensive understanding of its influence on mental health. As noted by de Lira & de Morais [53], resilience encompasses both individual (e.g., personal traits, learned capacity) and community resilience (e.g., family socialization, LGB community socialization). Future studies should address various types of resilience across different psychological pathways among individuals with multiple minority identities [67]. As an addition to the deficit mindset (which focuses on the symptoms and stressors endured by minority individuals), the field of public mental health should also examine the concept of resilience and encourage protective factors as related to minority individuals’ unique identity backgrounds. Despite the attenuating effect of inimical discrimination on depression by re, the focus on individual resilience does not diminish the pressing needs for fundamental changes to eradicate system-level discrimination or prejudice underlying minority stress. Our work once again highlights that the association between discriminatory experiences and adverse mental health outcomes is strong and lasting. This is another call for action that every aspect of the society, from public health policy-makers, mental health providers, to grassroot community groups, need to join efforts and foster a safer environment for minority individuals to thrive.

### Limitations

Results and interpretations of the present study should be considered in light of several limitations. Like all web-based studies, this sample consisted of voluntary participants who were younger, college educated, and had access to digital devices and internet. Therefore, results may not be generalizable for those who are older, less educated, and have limited access to Chinese LGBT communities in the U.S. Meanwhile, the cross-sectional design limits the establishment of causality between variables. Future longitudinal studies could investigate temporal relationships between acculturative stress, LGBT community connectedness, and mental health outcomes. Moreover, our study focused specifically on Chinese sexual minority individuals in the U.S., comparative studies on other ethnic and cultural groups could offer insights into how different cultural backgrounds impact the life experiences in sexual minority individuals across nations. Next, since we excluded gender minority individuals in the study; future studies should examine the intersectionality of gender minority individuals in term of acculturation, gender identity related experiences, access to medical services, and quality of life. Meanwhile, Yeo et al. [68] explained that discriminatory experiences in relation to self-concept may be differed for Asian/Asian American due to their country of origin. Asian Americans who were born and raised in the U.S. might have different emotional health when encounter discrimination compared to those who were born in other countries. Future studies could further examinate psychological wellbeing with its factors as acculturation, life experiences, immigration status within Asian/Chinese Americans. Last but not the least, the reliance on self-report measures for variables such as acculturative stress and mental health outcomes may be subject to response biases and potential social desirability effects. The inclusion of objective measures or multiple informants could strengthen the validity of the findings.

## Conclusion

Notwithstanding the limitations, the current study indicated that the relationship between discriminatory experiences and depressive symptoms can be mediated by internalized homophobia in Chinese sexual minority individuals residing in the U.S. (statistical significance for the role of internalized transphobia as a mediator and resilience as a moderator). Furthermore, the results revealed that resilience moderated the positive effect of internalized homophobia on depressive symptoms. The results showed the critical role of resilience in adjusting daily life encounter for individuals with intersecting identities in the U.S., providing evidence for further mental health intervention to Chinese sexual minority population.

## Data Availability

The data underlying the results presented in the study are available from the corresponding author by reasonable request.

## Acknowledgments

The authors extend their gratitude to the participants for their invaluable contributions and time. Sincere appreciation is extended to the Chinese Rainbow Network (CRN: https://www.crn.ngo) for their outstanding efforts in empowering Chinese LGBTQ^+^ individuals overseas. In addition, we extend our thanks to Huan Shen, Kyra Xu, Scoot, Hayley Huang, Tianqi Jiang, Jing Hong, Ethan Zhang, Alex, and other volunteers at CRN for their support in recruitment support and advocacy.

## References

1. Tessler H, Choi M, Kao GS. The anxiety of being Asian American: hate crimes and negative biases during the COVID-19 pandemic. Am J Crim Justice. 2020;45:636–646.

2. Pascoe EA, Richman SL. Perceived discrimination and health: a meta-analytic review. Psychol Bull. 2009;135(4):531–554. doi:10.1037/a0016059.

3. Florez E, Cohen K, Ferenczi N, Linnell K, Lloyd J, Goddard L, et al. Linking recent discrimination-related experiences and wellbeing via social cohesion and resilience. J Posit Psychol Wellbeing. 2020;4(1S):92–104.

4. Meyer IH. Prejudice, social stress, and mental health in lesbian, gay, and bisexual populations: conceptual issues and research evidence. Psychol Bull. 2003;129(5):674–697.

5. Pahl K, Wang J, Sanichar N, Williams S, Nick GA, Wang L, et al. Anti-Asian attitudes in the context of the COVID-19 pandemic: an exploratory study. J Racial Ethn Health Disparities. 2023;10(4):1947–1954. doi:10.1007/s40615-022-01376-6.

6. Bowleg L, Huang J, Brooks KD, Black AE, Burkholder G. Triple jeopardy and beyond: multiple minority stress and resilience among Black lesbians. J Lesbian Stud. 2003;7:108–137.

7. Torres L, Mata-Greve F, Bird C, Herrera Hernandez E. Intersectionality research within Latinx mental health: conceptual and methodological considerations. J Latina/o Psychol. 2018;6(4):304–317. doi: 10.1037/lat0000122.

8. Cyrus KD. Multiple minorities as multiply marginalized: applying the minority stress theory to LGBTQ people of color. J Gay Lesbian Ment Health. 2017;21:194–202. doi: 10.1080/19359705.2017.1320739.

9. Yette EM, Ahern J. Health-related quality of life among Black sexual minority women. Am J Prev Med. 2018;55(3):281–289. doi: 10.1016/j.amepre.2018.04.037.

10. Kudler BA. Confronting race and racism: social identity in African American gay men. Master’s thesis, Smith College School for Social Work. 2007. Available from: https://scholarworks.smith.edu/theses/1341/

11. Malebranche DJ, Fields EL, Bryant LO, Harper SR. Masculine socialization and sexual risk behaviors among Black men who have sex with men: a qualitative exploration. Men Masculinities. 2009;12(1):90–112. doi: 10.1177/1097184X07309504.

12. Diamond LM, Alley J. Rethinking minority stress: a social safety perspective on the health effects of stigma in sexually-diverse and gender-diverse populations. Neurosci Biobehav Rev. 2022;138:104720. doi: 10.1016/j.neubiorev.2022.104720.

13. Hatzenbuehler ML. How does sexual minority stigma “get under the skin”? A psychological mediation framework. Psychol Bull. 2009;135(5):707–730. doi: 10.1037/a0016441.

14. Shih M, Wilton LS, Does S, Goodale BM, Sanchez DT. Multiple racial identities as sources of psychological resilience. Soc Personal Psychol Compass. 2019;13(6):1–13. doi: 10.1111/spc3.12469.

15. Duran A. “Outsiders in a niche group”: using intersectionality to examine resilience for queer students of color. J Divers High Educ. 2021;14(2):217–227. doi: 10.1037/dhe0000144.

16. Njeze C, Bird-Naytowhow K, Pearl T, Hatala AR. Intersectionality of resilience: a strengths-based case study approach with indigenous youth in an urban Canadian context. Qual Health Res. 2020;30(13):2001–2018. doi: 10.1177/1049732320940702.

17. Ngamake ST, Walch SE, Raveepatarakul J. Discrimination and sexual minority mental health: mediation and moderation effects of coping. Psychol Sex Orientat Gend Divers. 2016;3(2):213–226. doi: 10.1037/sgd0000163.

18. Gee GC, Ro MJ, Rimoin AW. Seven reasons to care about racism and COVID-19 and seven things to do to stop it. Am J Public Health. 2020;110(7):954–955. doi: 10.2105/AJPH.2020.305712.

19. Woo B, Jun J. COVID-19 racial discrimination and depressive symptoms among Asian Americans: does communication about the incident matter? J Immigr Minor Health. 2022;24(1):78–85. doi: 10.1007/s10903-021-01167-x.

20. Liu SR, Davis EP, Palma AM, Stern HS, Sandman CA, Glynn LM. Experiences of COVID-19-related racism and impact on depression trajectories among racially/ethnically minoritized adolescents. J Adolesc Health. 2023;72(6):885–891. doi: 10.1016/j.jadohealth.2022.12.020.

21. Newcomb ME, Mustanski B. Internalized homophobia and internalizing mental health problems: a meta-analytic review. Clin Psychol Rev. 2010;30(8):1019–1029. doi: 10.1016/j.cpr.2010.07.003.

22. Brown AL, Matthews DD, Meanley S, Brennan-Ing M, Haberlen S, D’Souza G, et al. The effect of discrimination and resilience on depressive symptoms among middle-aged and older men who have sex with men. Stigma Health. 2022;7(1):113–121. doi: 10.1037/sah0000327.

23. Williams DR, Yu Y, Jackson JS, Anderson NB. Racial differences in physical and mental health: socio-economic status, stress and discrimination. J Health Psychol. 1997;2(3):335–351. doi: 10.1177/135910539700200305.

24. Fortuna LR, Porche MV, Alegria M. Political violence, psychosocial trauma, and the context of mental health services use among immigrant Latinos in the United States. Ethn Health. 2008;13(5):435–463. doi: 10.1080/13557850701837286.

25. John DA, De Castro AB, Martin DP, Duran B, Takeuchi DT. Does an immigrant health paradox exist among Asian Americans? Associations of nativity and occupational class with self-rated health and mental disorders. Soc Sci Med. 2012;75(12):2085–2098. doi: 10.1016/j.socscimed.2012.01.035.

26. Chan KTK, Tran TV, Nguyen TN. Cross-cultural equivalence of a measure of perceived discrimination between Chinese-Americans and Vietnamese-Americans. J Ethn Cult Divers Soc Work. 2012;21(1):20–36. doi: 10.1080/15313204.2011.647348.

27. Nicholson HL. Associations between major and everyday discrimination and self-rated health among US Asians and Asian Americans. J Racial Ethn Health Disparities. 2020;7:262–268. doi: 10.1007/s40615-019-00654-0.

28. Wagner GJ. Internalized homophobia scale. In: Fisher TD, Davis CM, Yarber WL, eds. Handbook of sexuality-related measures. Routledge; 1998:371–372.

29. McLaren S, Castillo P. The relationship between a sense of belonging to the LGBTIQ+ community, internalized heterosexism, and depressive symptoms among bisexual and lesbian women. J Bisexuality. 2021;21(1):1–23. doi: 10.1080/00918369.2015.1083779.

30. Quinn KG, Kelly JA, DiFranceisco WJ, Tarima SS, Petroll AE, Sanders C, et al. The health and sociocultural correlates of AIDS genocidal beliefs and medical mistrust among African American MSM. AIDS Behav. 2018;22:1814–1825. doi: 10.1007/s10461-016-1657-6.

31. Connor KM, Davidson JR. Development of a new resilience scale: The Connor-Davidson resilience scale (CD-RISC). Depress Anxiety. 2003;18(2):76–82. doi: 10.1002/da.10113.

32. Cheng C, Dong D, He J, Zhong X, Yao S. Psychometric properties of the 10-item Connor-Davidson Resilience Scale (CD-RISC-10) in Chinese undergraduates and depressive patients. J Affect Disord. 2020;261:211–220. doi: 10.1016/j.jad.2019.10.018.

33. Wang Y, Lao CK, Wang Q, Zhou G. The impact of sexual minority stigma on depression: the roles of resilience and family support. Sex Res Social Policy. 2022;19:442–452. doi: 10.1007/s13178-021-00558-x.

34. Radloff LS. The CES-D scale: a self-report depression scale for research in the general population. Appl Psychol Meas. 1977;1(3):385–401. doi: 10.1177/014662167700100306.

35. Yan H, Li X, Li J, Wang W, Yang Y, Yao X, et al. Association between perceived HIV stigma, social support, resilience, self-esteem, and depressive symptoms among HIV-positive men who have sex with men (MSM) in Nanjing, China. AIDS Care. 2019;31(9):1069–1076. doi: 10.1080/09540121.2019.1601677.

36. Zhao M, Xiao D, Wang W, Wu R, Dewaele A, Zhang W, et al. Association of sexual minority status, gender nonconformity with childhood victimization and adulthood depressive symptoms: a path analysis. Child Abuse Negl. 2021;111:104822. doi: 10.1016/j.chiabu.2020.104822.

37. JASP Team. JASP. Version 0.18.1[software]. Available at: https://jasp-stats.org/download/

38. Rosseel Y. Lavaan: An R package for structural equation modeling and more. Version 0.5-12 (BETA). J Stat Softw. 2012; 48(2):1–36.

39. Biesanz JC, Falk CF, Savalei V. Assessing mediational models: testing and interval estimation for indirect effects. Multivariate Behav Res. 2010;45(4):661–701. doi: 10.1080/00273171.2010.498239.

40. Mallory AB, Russell ST. Intersections of racial discrimination and LGB victimization for mental health: a prospective study of sexual minority youth of color. J Youth Adolesc. 2021;50(7):1353–1368. doi: 10.1007/s10964-021-01443-x.

41. Sutter M, Perrin PB. Discrimination, mental health, and suicidal ideation among LGBTQ people of color. J Couns Psychol. 2016;63(1):98–105. doi: 10.1037/cou0000126.

42. Liang Z, Huang YT. “Strong together”: minority stress, internalized homophobia, relationship satisfaction, and depressive symptoms among Taiwanese young gay men. J Sex Res. 2022;59(5):621–631. doi: 10.1080/00224499.2021.1947954.

43. Wei M, Heppner PP, Ku TY, Liao KYH. Racial discrimination stress, coping, and depressive symptoms among Asian Americans: a moderation analysis. Asian Am J Psychol. 2010;1(2):136–150. doi: 10.1037/a0020157.

44. Hswen Y, Xu X, Hing A, Hawkins JB, Brownstein JS, Gee GC. Association of “# covid19” versus “# chinesevirus” with anti-Asian sentiments on Twitter: March 9–23, 2020. Am J Public Health. 2021;111(5):956–964. doi: 10.2105/AJPH.2021.306154.

45. Li M. Discrimination and psychiatric disorder among Asian American immigrants: a national analysis by subgroups. J Immigr Minor Health. 2014;16(6):1157–1166. doi: 10.1007/s10903-013-9920-7.

46. Cheah CS, Wang C, Ren H, Zong X, Cho HS, Xue X. COVID-19 racism and mental health in Chinese American families. Pediatrics. 2020;146(5). doi: 10.1542/peds.2020-021816

47. Wu C, Qian Y, Wilkes R. Anti-Asian discrimination and the Asian-white mental health gap during COVID-19. Ethn Racial Stud. 2021;44(5):819–835.

48. Chiu C. Sexual racism and mental health among Asian/Asian American sexual minority men. Dissertation, University of Massachusetts Boston. 2023. Available from: https://scholarworks.umb.edu/doctoral_dissertations/874/

49. Choi AY, Israel T. Centralizing the psychology of sexual minority Asian and Pacific Islander Americans. Psychol Sex Orientat Gend Divers. 2016;3(3):345–356. doi: 10.1037/sgd0000184.

50. Sue DW, Capodilupo CM, Torino GC, Bucceri JM, Holder AMB, Nadal KL, et al. Racial microaggressions in everyday life: implications for clinical practice. Am Psychol. 2007;62(4):271–286. doi: 10.1037/0003-066X.62.4.271.

51. Sue DW. Microaggressions, marginality, and oppression: an introduction. In: Sue DW, editor. Microaggressions and marginality: Manifestation, dynamics, and impact. John Wiley & Sons, Inc.; 2010. p.3–22.

52. Luthar SS, Ebbert AM, Kumar NL. Risk and resilience among Asian American youth: ramifications of discrimination and low authenticity in self-presentations. Am Psychol. 2021;76(4):643–656. doi: 10.1037/amp0000764.

53. de Lira AN, de Morais NA. Resilience in lesbian, gay, and bisexual (LGB) populations: an integrative literature review. Sex Res Soc Policy. 2018;15(3):272–282.

54. Feinstein BA, Goldfried MR, Davila J. The relationship between experiences of discrimination and mental health among lesbians and gay men: an examination of internalized homonegativity and rejection sensitivity as potential mechanisms. J Consult Clin Psychol. 2012;80(5):917.

55. Puckett JA, Newcomb ME, Garofalo R, Mustanski B. The impact of victimization and neuroticism on mental health in young men who have sex with men: internalized homophobia as an underlying mechanism. Sex Res Soc Policy. 2016;13(3):193–201. doi: 10.1007/s13178-016-0239-8.

56. Sun S, Hoyt WT, Tarantino N, Pachankis JE, Whiteley L, Operario D, et al. Cultural context matters: testing the minority stress model among Chinese sexual minority men. J Couns Psychol. doi: 10.1037/cou0000535.

57. Morrow DF. Older gays and lesbians. J Gay Lesbian Soc Serv. 2001;13(1-2):151–169. doi: 10.1300/J041v13n01_11.

58. Cui L. “I had to get married to protect myself”: gay academics’ experiences of managing sexual identity in China. Asian J Soc Sci. 2022;50(4):260–267. doi: 10.1016/j.ajss.2022.05.007.

59. Song C, Xie H, Alizai A, Chatterjee JS. “I did not know I was gay”: sexual identity development and fluidity among married tongzhi in China. Cult Health Sex. 2022;24(12):1681–1694. doi: 10.1080/13691058.2021.1996631.

60. Sung MR, Szymanski DM, Henrichs-Beck C. Challenges, coping, and benefits of being an Asian American lesbian or bisexual woman. Psychol Sex Orientat Gend Divers. 2015;2(1):52–64. doi: 10.1037/sgd0000085.

61. Ching THW, Lee SY, Chen J, So RP, Williams MT. A model of intersectional stress and trauma in Asian American sexual and gender minorities. Psychol Violence. 2018;8(6):657–668. doi: 10.1037/vio0000204.

62. Scandurra C, Bochicchio V, Amodeo AL, Esposito C, Valerio P, Maldonato NM, et al. Internalized transphobia, resilience, and mental health: applying the Psychological Mediation Framework to Italian transgender individuals. Int J Environ Res Public Health. 2018;15(3):508. doi: 10.3390/ijerph15030508.

63. Li X, Yan H, Wang W, Yang H, Li S. Association between enacted stigma, internalized stigma, resilience, and depressive symptoms among young men who have sex with men in China: a moderated mediation model analysis. Ann Epidemiol. 2021;56:1–8.

64. Meyer IH. Resilience in the study of minority stress and health of sexual and gender minorities. Psychol Sex Orientat Gend Divers. 2015;2(3):209.

65. Schnarrs PW, Stone AL, Salcido R, Georgiou C, Zhou X, Nemeroff CB. The moderating effect of resilience on the relationship between adverse childhood experiences (ACEs) and quality of physical and mental health among adult sexual and gender minorities. Behav Med. 2020;46(3-4):366–374. doi: 10.1080/08964289.2020.1727406.

66. Doan SN, Yu SH, Wright B, Fung J, Saleem F, Lau AS. Resilience and family socialization processes in ethnic minority youth: illuminating the achievement-health paradox. Clin Child Fam Psychol Rev. 2022;25(1):75–92. doi: 10.1007/s10567-022-00389-1.

67. Pharr JR, Chien LC, Gakh M, Flatt JD, Kittle K, Terry E. Moderating effect of community and individual resilience on structural stigma and suicidal ideation among sexual and gender minority adults in the United States. Int J Environ Res Public Health. 2022;19(21):14526. doi: 10.3390/ijerph192114526.

68. Yeo AJ, Halpern LF, Flagg AM, Lin B. Discrimination and depressive symptoms among Black and Asian American college students: shared and group-specific processes of self-concept. Cult Divers Ethn Minor Psychol.

